# SARS-CoV-2 evolution and evasion from multiple antibody treatments in a cancer patient

**DOI:** 10.1101/2022.06.25.22276445

**Authors:** Guy Shapira, Chen Weiner, Reut Sorek Abramovich, Odit Gutwein, Nir Rainy, Patricia Benveniste-Levkovitz, Ezra Gordon, Adina Bar Chaim, Noam Shomron

## Abstract

Infection with severe acute respiratory syndrome coronavirus 2 (SARS-CoV-2) in immunocompromised patients may lead to accelerated viral mutation rate, immune evasion and persistent viral shedding over many months. Here we report the case of a severely immunocompromised cancer patient infected with the Delta variant of SARS-CoV-2 for over 8 months. Genome sequencing of samples taken after repeated monoclonal antibody treatments reveal the emergence and accumulation of mutations enabling escape from neutralization by antibodies. Mutations emerging in accessory and non-structural viral proteins target specific residues of immunomodulatory domains, potentially leading to loss of some functions, while preserving others. The mutated virus managed to completely overcome neutralization by monoclonal antibodies while remaining viable and infective. Our results suggest that the loss of specific immunomodulatory viral functions might confer a selective advantage in immunocompromised hosts. We also compare between mutations emerging in the presence and absence of neutralizing antibodies.

**Highlights:**

- SARS-CoV-2 undergoes rapid evolution in an immunocompromised, chronically infected cancer patient, overcoming neutralization by two monoclonal antibody cocktail treatments
- Receptor binding domain (RBD) mutations emerging after monoclonal antibody treatment enable effective escape from neutralization in the absence of adaptive immunity
- Some emerging mutations are predicted to disrupt immunomodulatory viral proteins, including prevention of ORF8 homodimerization, mis-localization of ORF3a in host cells and alteration of the host-suppressive function of NSP1

## Introduction

The evolution of severe acute respiratory syndrome coronavirus 2 (SARS-CoV-2) in healthy human hosts is extremely limited, as reflected by its low overall genetic diversity^1^. The duration in which SARS-CoV-2 is transmissible is usually limited to less than 14 days^2^ and most emerging mutations are not transmitted due to strong purifying selection^3^. In contrast, when SARS-CoV-2 infects an immunocompromised individual, the virus can overcome the host’s impaired immunity, remaining infectious for many months and accumulating unusual mutations^4^. The risk of persistent SARS-CoV-2 infection varies according to the type of immunosuppression therapy, with recipients of B-cell-depleting therapy, such as the anti-CD20 medication rituximab, being at exceptionally high risk^5^. Some antiviral therapies might be used to compensate for the impaired immunity of such patients, including monoclonal or polyclonal exogenous antibodies, ivermectin and remdesivir. However, the success of these treatments is not always guaranteed, due to resistant viral mutants that emerge in some cases^6–9^.

The increased evolutionary capacity of SARS-CoV-2 in immunocompromised patients is also a threat to public health, giving rise to mutations that increase viral fitness and enhance escape from neutralizing antibodies^9^. It is hypothesized that such cases of persistent infection are at least partly responsible for the emergence of novel variants of concern (VOC), causing global periodic surges of infection^4^.

Here we present to you the case of a female cancer patient in her 60s with multiple malignant diseases, who was diagnosed with SARS-CoV-2 in September 2021 and remains infected as of May 2022.

## Results

### Clinical presentation of the chronically infected patient

A female patient aged 60-69 was treated starting in June 2021 at Shamir Medical Center hospital, Be’er Yaacov, Israel as an oncological, immunocompromised patient (complete clinical timeline in Table 1). Oncological findings included: Malignant melanoma, Diffuse large B-cell lymphoma (DLBCL) and Squamous Cell Carcinoma (SCC). Background diseases included: fibromyalgia, treated asthma and Chronic Obstructive Pulmonary Disease (COPD). The patient was identified as Covid-19 positive during a routine visit in September 2021, and since then has tested positive 15 times and borderline positive once. The patient mostly presented as asymptomatic. Three Nasopharyngeal samples were obtained over the duration of infection and sent to viral whole-genome sequencing (Table 1).

**Table 1:** A clinical timeline of sample collection, assay results and other treatments with relevance to COVID-19 and immunity. Dates are given in a generalized Month-Year format and list in order of occurrence, from least to most recent.

| Date | Description |
| --- | --- |
| June 2021 | Initial Melanoma findings |
| September 2021 | First positive SARS-CoV-2 PCR result (17Ct S gene) |
| September 2021 | First REGEN-COV monoclonal treatment (casirivimab and imdevimab) |
| October 2021 | Rituximab treatment (part of the R-CHOP protocol) |
| October 2021 | Two doses of R-CHOP chemotherapy |
| November 2021 | R-CHOP chemotherapy |
| December 2021 | R-CHOP chemotherapy |
| December 2021 | Collection of sample S1 (Positive PCR result) |
| December 2021 | Second REGEN-COV monoclonal treatment (casirivimab and imdevimab) |
| December 2021 | Positive SARS-CoV-2 PCR result (27Ct S gene) |
| December 2021 | Collection of sample S2 (19Ct S gene) |
| December 2021 | SARS-CoV-2 trimeric S IgG serology result: 5520 (AU/ml) |
| January 2022 | R-CHOP chemotherapy and Keytruda treatment |
| January 2022 | Last Rituximab treatment (part of the R-CHOP protocol) |
| February 2022 | Keytruda treatment |
| February 2022 | Positive result from infectivity assay |
| February 2022 | Positive SARS-CoV-2 PCR result (25Ct S gene) |
| March 2022 | Borderline-positive SARS-CoV-2 PCR results (33Ct and 35Ct S gene) |
| May 2022 | Collection of sample S3 (19Ct S gene) |

### Origin of the SARS-CoV-2 infection and accumulating mutations

Whole-genome sequence analysis identified the SARS-CoV-2 genome as belonging to the Delta variant, lineage AY.43, genetically related to contemporary viral genomes collected in Israel and across Europe (Figure 1). To identify mutations that emerged within the immunocompromised host over the course of infection, we compared the mutational profile of the viral samples, as well as that of the sample representing the closest phylogenetic relative (Figure 2). The earliest sample (sample S1) is substantially more mutated than its closest phylogenetic neighbor, with 10 new substitutions. Genomes sequenced from samples acquired later in the course of infection show a pattern of accumulating mutation, with 7 new mutations in less than a month and another 10 mutations roughly 3 months later.

**Figure 1:**
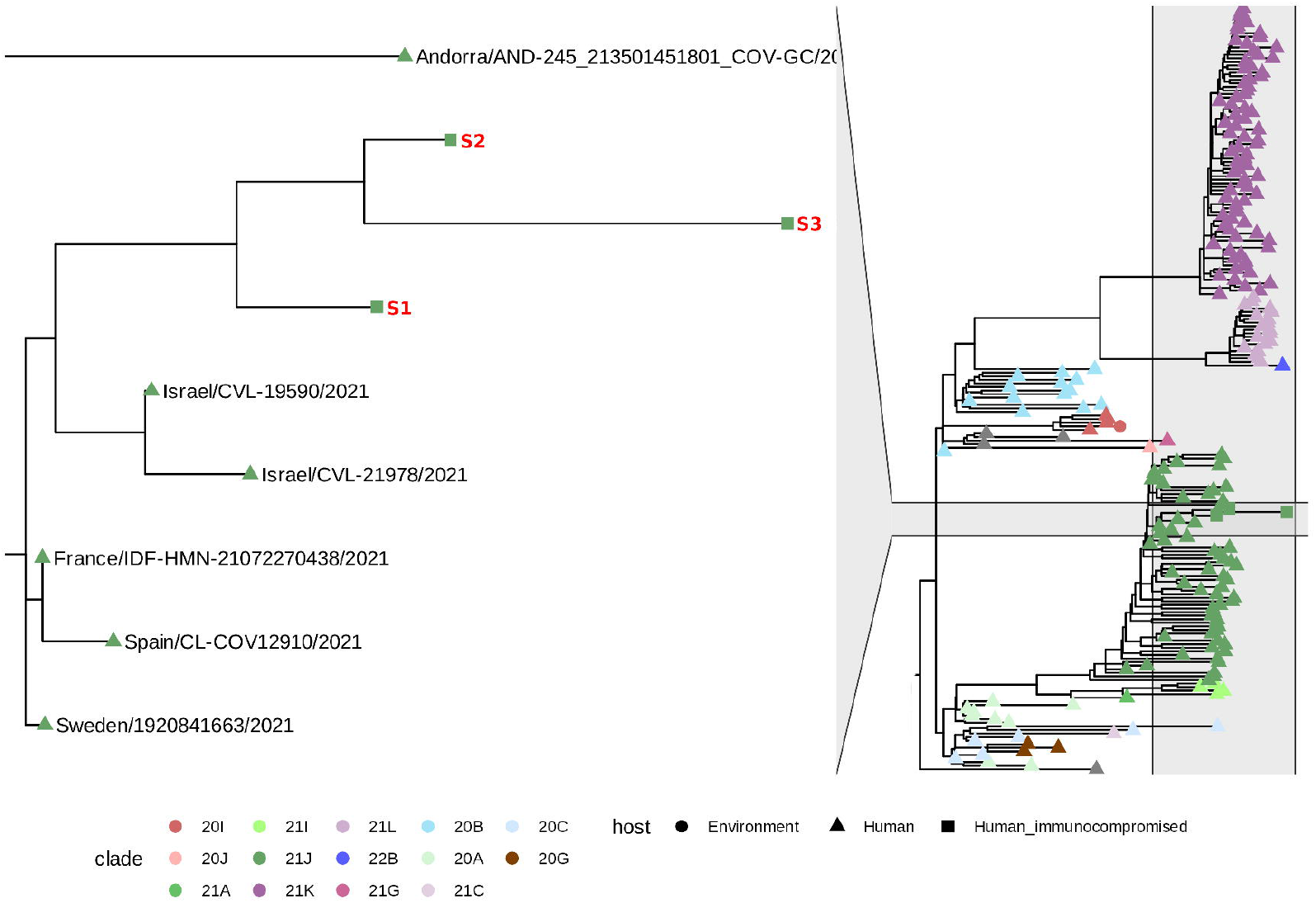
Time resolved phylogeny of SARS-CoV-2 genomes in a global context (right) and in the context of samples obtained from the immunocompromised patient (left). Samples from the immunocompromised patient are marked with squares and bold red labels.

**Figure 2:**
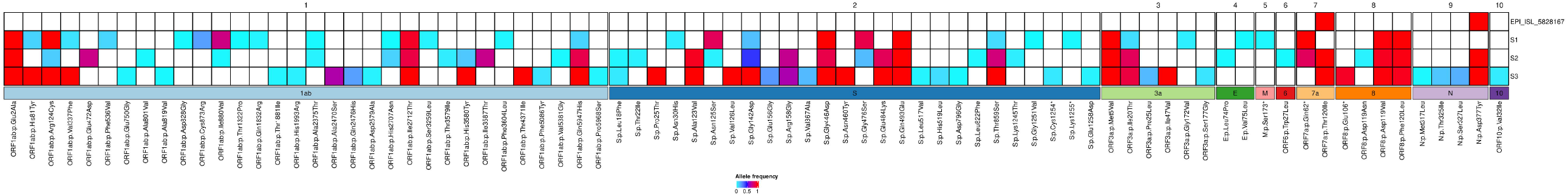
Protein-altering mutations in the viral sequences sampled from the immunosuppressed patient (only mutations that differ from the closest ancestral sequence are shown). The patient-derived viral samples are ordered from the earliest to the most recent (S1, S2 and S3). Cells are colored according to the mutation allele frequency (fraction of mutated reads) and annotated with the name of the overlapping gene.

To better capture the viral variation within the host, we include lower frequency mutations that were called with high confidence (see methods). We distinguish between minor mutations, representing a minority of the viral intra-host population (Allele frequency (AF) < 0.5) from the major mutations that became fixed (AF > 0.5).

The distribution of emerging mutations is uneven, with the accessory proteins, ORF8, ORF10 and ORF3a, accumulating the most mutations (10.9, 8.6 and 6 mutations per kilobase (mKb), respectively). The Spike gene is the most mutated structural protein coding gene, with 23 distinct nonsynonymous mutations, 8 of which overlap the locus coding for the receptor binding domain (RBD), making it the most densely mutated locus overall (∼6 mKb in the Spike gene, ∼12 in the RBD alone).

### Chronology of antiviral treatment and emerging Spike gene mutations

The patient was initially diagnosed with SARS-CoV-2 infection in September, 2021, with high viral load (17Ct), characteristic of Delta variant infections from this period^10^. Within the same month, the patient was treated with the monoclonal antibody cocktail REGEN-COV (casirivimab and imdevimab). Following the monoclonal antibody treatment, two negative PCR test results indicated the infection was resolved and cancer therapy was initiated. Over the two following months, the patient received immunochemotherapy, according to the R-CHOP protocol, including the immunosuppressant Rituximab (Anti-CD20). In December, 2021, a positive PCR test indicated that the infection persisted and the first viral genome sample was sequenced (sample S1).

Sample S1 harbors 3 RBD mutations: G446D, G476S and Q493E (Figure 1). Additionally, 3 missense mutations emerged in the N terminal domain of the Spike gene and an additional 3 minor mutations emerged in the S2 subunit of the spike protein. On December 2021, the patient received another dose of REGEN-COV, which again failed to fully resolve the infection and was followed by an increase in viral load, despite high concentrations of serum antibodies (5520 AU/ml)(Table 1).

In sample S2, obtained less than a month after the second REGEN-COV treatment, we find multiple newly emerged mutations, including 4 spike mutations of minor frequencies and the major RBD mutation E484K.

The latest viral genome sample (sample S3), collected May 2022, has 10 newly emergent Spike mutations. The S3 genome has 4 new RBD mutations, one major (N460Y) and three minor (V367A, L517V and H519L). Interestingly, the RBD mutation G476S, which was first observed in sample S1, decreased in frequency in sample S2 and was completely absent from sample S3.

### The impact of emerging mutations on antibody recognition

We see an accumulation of RBD mutations over time, specifically in amino acids found to enable escape from binding by antibodies (Figure 3). The mutations G446D, E484K and Q493E all confer escape from the REGEN-COV monoclonal antibodies used to treat the patient, according to deep mutational scanning^11^.

**Figure 3:**
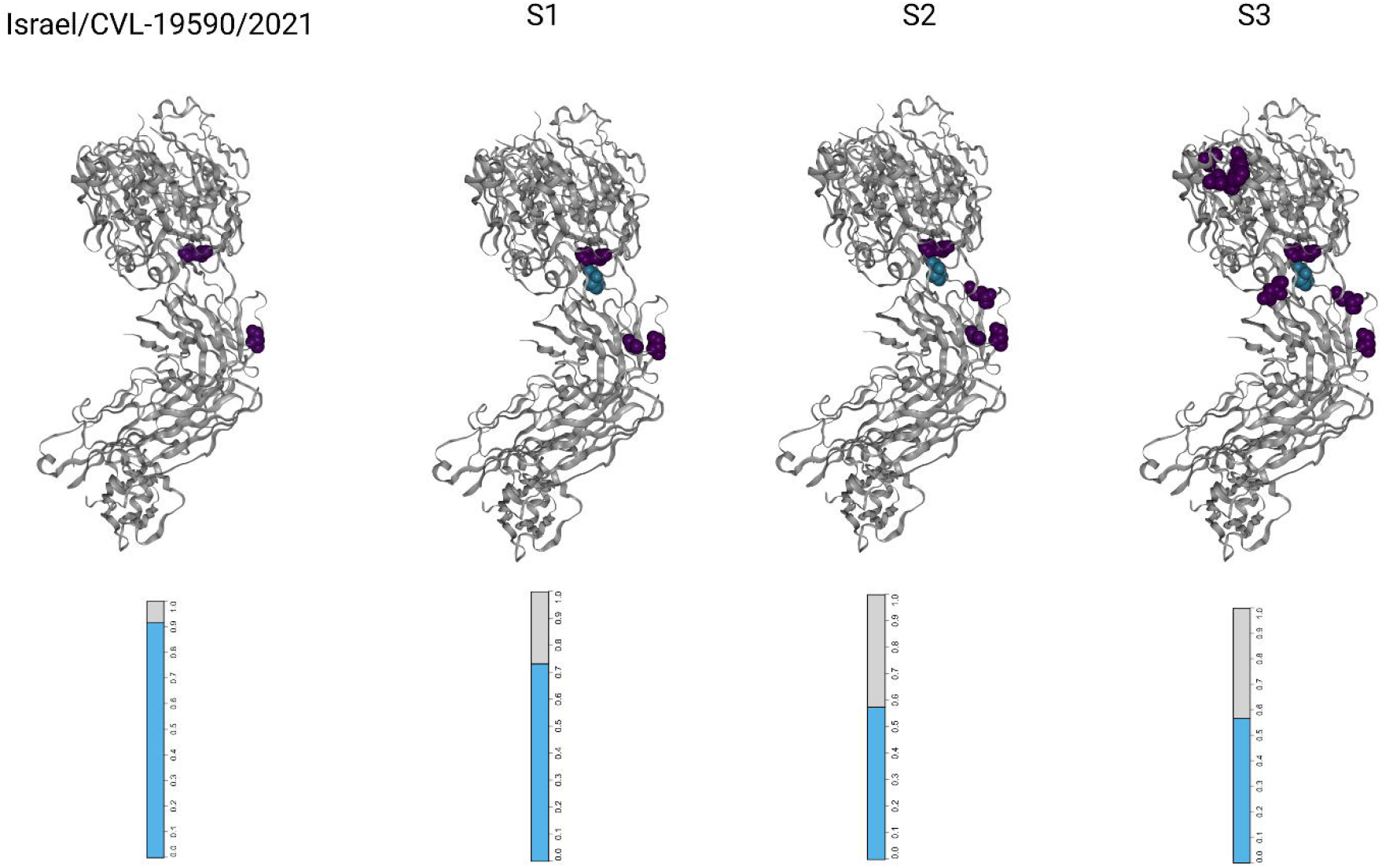
Visualizations of the SARS-CoV-2 receptor binding domains (RBDs), marked according to altered amino acid residues found in each sample. Each mutated residue is colored according to the magnitude of the largest-effect escape mutation at each site, as measured for the REGN10933+REGN10987 monoclonal antibody cocktail (see methods). The bar-plots at the bottom represent the estimated fraction antibodies that were either bound (cyan) or escaped (gray) from neutralization by polyclonal antibodies for each sample (see methods).

E484K is significant for being an example of convergent evolution, evolving independently in multiple VOCs and conferring extensive escape from binding by multiple antibodies^12^. The allelic fraction of E484K increased from 85% in sample S2 to nearly 100% in S3, possibly indicating positive selection.

The 4 RBD mutations that emerged in sample S3, over 5 months since REGEN-COV administration, are the least influential on antibody binding and, with the exception of the major mutation N460Y, they account for relatively small fractions of the viral population (2%-15%).

## Discussion

### Limited humoral response and monoclonal antibodies effectively evaded by SARS-CoV-2 RBD mutations

In this case study, we characterize the evolution of SARS-CoV-2 in an immunocompromised cancer patient, covering 6 out of 9 months of infection (the infection remains unresolved at the time of writing). Over the long course of infection, the virus survived two treatments of potent monoclonal antibody therapy, remaining infectious and viable despite an exceptionally high concentration of serum antibodies^13^. The inability of adaptive immunity to mitigate the infection could be partly attributed to treatment with Rituximab, an anti-CD20 immunosuppressant known to severely impair serological response to SARS-CoV-2^5^.

According to prior studies, it is likely that the full extent of humoral antibodies is composed solely of endogenous antibodies from the first month of infection and the exogenous REGEN-COV antibodies^14^. Following these assumptions, we hypothesize that the mutations acquired before collection of sample S1 are sufficient to escape effective neutralization by both the REGEN-COV monoclonal cocktail antibodies and the endogenous antibodies produced before treatment with Rituximab. The second REGEN-COV monoclonal antibody treatment was accompanied by accumulation of additional RBD mutations, enabling greater escape from antibody binding (Figure 3) and likely decreasing the efficacy of the treatment even further. Following the second antibody treatment, viral shedding remained persistent and infective, as evident by the positive infectivity assay taken roughly two months later. While mutations continued accumulating for another 5 months, none of the emerging RBD mutations were associated with increased escape from binding by either monoclonal or polyclonal mutations (Figure 3). The final Rituximab treatment was administered over four months before collection of sample S3, however, since the suppressive effects of anti-CD20 therapy typically last for over a year^14^, it is likely that humoral response remained negligible.

While repeated REGEN-COV treatments failed to achieve viral clearance, reports of similar cases suggest that co-administration with the antiviral drug Remdesivir might increase the odds of successfully resolving the infection^7^.

### Accelerated mutational rate of the Spike protein and escape from neutralizing antibodies

Consistent with previous studies, the immunocompromised viral genome had more intra-host mutations, greater enrichment of nonsynonymous mutations in the Spike protein and increased fixation of minor mutations when compared to samples taken from immunocompetent hosts^3^. Many of the emerging RBD mutations have been previously associated with escape from binding by monoclonal antibody treatment, implying that these mutations were partly responsible for the virus overcoming neutralization by antibodies and, through a process of strong positive selection, their emergence was a consequence of the antibody treatment^15^ (Figure 3).

Some RBD mutations, such as E484K, evolved independently in other immunocompromised individuals and separate SARS-CoV-2 lineages, an example of convergent evolution^12^. Spike protein amino acid residues that form antibody recognition sites, such as 446, 484 and 493, are frequently mutated in immunocompromised hosts to escape neutralization by a broad range of antibodies^16^.

Samples S1 and S2 were taken after treatment with the REGEN-COV monoclonal cocktail and contain a large number of previously described mutations that enable escape from binding by these specific antibodies. Results from deep mutational scanning experiments support the hypothesis that RBD mutations emerged in samples S1 and S2 primarily in response to selective pressure applied by the REGEN-COV antibodies. RBD mutations from sample S3, taken 5 months after the last REGEN-COV treatment, were not associated with improved escape from monoclonal or polyclonal antibodies. The extremely rare N460Y Spike mutation in sample S3 (200∼ occurrences in GISAID) might not be explained by selective pressure, but the fact that it is fixed (100% of viral reads) implies significant fitness advantage.

### Mutations in immunomodulatory viral proteins and their predicted impact on specific functional domains

Alongside Spike protein mutation, a broad distribution of mutations was found affecting accessory and non-structural proteins.

ORF3a, a large accessory protein with various immunomodulatory functions acting on the host^17^, acquired the rare signal peptide mutation M5V in sample S1 (<350 occurrences in GISAID) that remained fixed throughout. Closely downstream, the N-terminal ectodomain mutation I20T emerged in sample S1 and became fixed over the course of infection (7%, 84% and 90%, in samples S1,S2 and S3, respectively), implying it was subjected to positive selection. I47V in the transmembrane domain emerged as a major ORF3a mutation in sample S3, affecting one of the 6 residues involved in the formation of a hydrophobic seal required for the ion channel function of ORF3a^18^. The amino acid residues altered in the majority of the viral population are primarily associated with cellular localization functions of ORF3a^19^. Their relative rarities, combined with the predicted deleteriousness of neighboring mutations^20^, imply significant loss of function.

ORF8 is another accessory protein known to modulate host immunity factors^21^. Its mutations are specific to amino acid 119, including the major persistent D119V and the minor D119N, only in sample S2, closely followed by the major persistent F120L. These mutations specifically alter homodimeric interfaces of the ORF8 protein, with D119 forming a salt bridge and F120 forming a hydrogen bond^22^. In sample S3, the stop gain mutation E106* truncates 15 amino acids, including those affected by the substitutions. The specificity of the mutations to amino acids involved in the ORF8 homodimer formation suggest some benefit to its disruption, culminating in the complete truncating of the domain. Recent studies revealed a myriad of functions facilitated by ORF8, with implications for transmission dynamics, host immunity and male fertility. However, whether or not these functions depend on the dimeric form of the protein is yet to be determined^23–25^.

NSP1 is a non-structural protein that causes a reduction in translation of cellular transcripts, promotes degradation of host mRNAs and inhibits export of nuclear mRNA, inhibiting host IFN-I response^26^. Residues 1-117 and 122-130 of NSP1, which mediate these host-suppressive functions^27^ are affected by the mutations G2A, H81C and R124C, fixed in sample S3. It has been reported that the induction of NSP1 mutations in the same locus as R124C (R124A and K125A) prevents NSP1 from accelerating the degradation of host mRNA and decreases its ribosome binding, while increasing its suppression of both host and viral mRNA translation^28^. Based on these observations, we estimate that at least one of the three NSP1 mutations significantly alters host-pathogen interaction; this alteration is likely beneficial, since all three mutations are fixed in sample S3.

Other mutations that emerged in NSP2, NSP3, NSP10 and NSP14 became fixed over time, suggesting a selective advantage. However, their specific importance could not be inferred from the existing literature. Major deletions and loss of function mutations in accessory proteins are not uncommon^29^. Such mutations have been documented in immunocompromised patients and are generally associated with milder symptoms^30^. The accessory protein mutations emerging in this case are specific to residues of functional domains, implying a more nuanced effect, rather than more general loss of function mutations. The possible phenotypic consequences of such mutations are exemplified by NSP1, where the experimental alteration of similar residues led to a loss of mRNA degrading function, even though its translation suppressing function remained mostly unchanged^26^.

## Materials and methods

### RNA Extraction

RNA was extracted from nasopharyngeal swabs using a Biomek i7 automatic liquid handler (Beckman Coulter) by magnetic bead separation (RNAdvance Viral XP Reagent Kit, Beckman Coulter, C59543) according to the manufacturer’s instructions. Briefly, 200ul of sample was taken directly from the testing tube and added to a 2ml tube containing 150ul of lysis buffer (Beckman Coulter) in a BSL2 Biological Hood. Samples were kept at RT for 20 minutes to allow viral inactivation and proper lysis and subsequently were loaded to the liquid handler for reformatting into a 96-wells deep-well plate. 350ul of magnetic beads (RNAdvance Viral Bind-VBE) were added to each sample. Thereafter, the plate was incubated for 5 minutes to allow the binding of the magnetic beads to the RNA. After binding, the plate was automatically transferred to an on-deck 96-well magnet (Magnum FLX®, Alpaqua) for 5 min to allow bead settling. Samples were washed twice with 80% ethanol (Biolabls, Israel) and eluted in molecular grade water (Biolabls, Israel). Purified RNA was kept in a 96-wells format plate at -80□.

### Library Preparation and Sequencing

SARS-CoV-2 whole genome libraries were prepared using Illumina COVIDSeq protocol according to the manufacturer’s protocol (Illumina Inc, USA). The first strand synthesis was carried out on RNA samples isolated using RNAdvance Viral XP Reagent Kit. The synthesized cDNA was amplified using a multiplex polymerase chain reaction (PCR) protocol, producing 98 amplicons across the SARS-CoV-2 genome (https://artic.network/). The PCR amplified product was later processed for tagmentation and adapter ligation using IDT for Illumina Nextera UD Indexes Set A, B, C, D (384 indexes, 384 samples). Further enrichment and cleanup was performed as per protocols provided by the manufacturer (Illumina Inc). All samples were processed as batches in a 384-well plate that consisted of one of COVIDSeq positive control HT (CPC HT), two no template control (NTC) and one Negative Sample. Finally, these 384 libraries were pooled together into 8 pools of 48 Samples each. Pooled samples were quantified by Qubit 4.0 fluorometer using HS DS DNA kit (Invitrogen Inc.) and fragment sizes were determined by TapeStation 4150 via DNA HS D1000 kit (Agilent). The pooled libraries were further normalized to 4nM concentration and 5 μl of each normalized pool were combined in a new microcentrifuge tube. For sequencing, pooled libraries were denatured and neutralized with 0.2N NaOH and 400mM Tris-HCL (pH-8). Dual indexed paired-end sequencing with 149bp read length was carried out on NextSeq 550 platform (Illumina Inc).

### RT-PCR Viral Detection

Extracted RNA (5ul) was transferred to a 96 well PCR plate containing 20μl of TaqPathTM 1-step Multiplex Master Mix No ROX (Applied Bioscience). Followed by one-step RT-PCR (Thermo-Fisher TaqPath COVID-19 assay kit). Thereafter, the plate was sealed with MicroAmp clear adhesive strip (Applied Bioscience). The plate was then loaded onto a QuantStudio™ 5 Real Time PCR System (Applied Bioscience) and the following amplification program was used: 25°C for 2 minutes, X1 cycle 53°C for 10 minutes, X1 cycle 95°C for 2 minutes, X1 cycle 95°C for 3 seconds, followed by 60°C for 30 seconds, X40 cycles Ct threshold values were preset using the following values/parameters: MS2-15,000; by cycle 37; S gene-20,000 by cycle 37; Orf1ab-20,000 by cycle 37; Ngene-20,000 by cycle 37. Samples that passed the Threshold is a Ct value >37 were re-tested or considered weak positive.

### Serological Viral Detection

The LIAISON® SARS-CoV-2 TrimericS IgG assay was in use starting March 2021 till present, all tests were performed on the LIAISON® SARS-CoV-2 S1/S2 IgG (DiaSorin, Saluggia, Italy): A chemiluminescent immunoassay (CLIA) for quantitative determination of anti-S1 and anti-S2 specific IgG antibodies using magnetic beads coated with S1 and S2 antigens. SARS-CoV-2 S1/S2 IgG antibody concentrations are automatically calculated and expressed as arbitrary units (AU/mL), with a positive cutoff level of 15.0 AU/mL (according to manufacturer declaration diagnostic sensitivity above 15 days of symptoms onset is 97.4% and specificity is 98.9%).

### Bioinformatic analysis, annotations and modeling

Raw reads were trimmed and filtered using fastp v0.22.0^31^, then aligned to the MN.908947 reference assembly using bwa-mem 2.2.1^32^. samtools v1.12 was used for primer trimming and variant calling, with the latter performed by a combination of samtools mpileup and iVar 1.3.1^33^. To remove low quality variant calls, we performed the process independently for each of the four sequencing lanes, including only variants that passed the significance threshold in multiple lanes. The phylogenetic tree was constructed using the Nexstrain v11.0 pipeline^34^, with the global nextregions genomes as the context (updated 9/5/2022) and most similar genome sequences obtained from UShER with full GISAID data^35^. Variants were annotated using SnpEff v5.1^36^. Assessment of antibody escape was calculated using measurements from deep mutational scanning experiments^11^.The global prevalence of various mutations was estimated using statistics from the GISAID database^37^ (updated 12/6/2022).

Estimation of the effect of individual RBD mutations on antibody binding was obtained from the deep mutational scanning experiments conducted by Starr et al.^11^ and visualized using dms-view (https://jbloomlab.github.io/SARS-CoV-2-RBD_MAP_clinical_Abs) with “max escape” scores according to binding by REGN10933+REGN10987 monoclonal antibodies. The combined effect of RBD mutations on binding by polyclonal antibodies was estimated using the escape calculator (https://jbloomlab.github.io/SARS2_RBD_Ab_escape_maps/escape-calc)^15^, set for antibodies elicited by pre-Omicron SARS-CoV-2.

## Data Availability

Full sequencing data and reproducible analysis code will be released upon publishing.

## Ethics declarations

The Ethics Committee of Shamir Medical Center has approved this study. Patient Data cannot be shared publicly due to patient confidentiality. Data are available from the Shamir Medical Center Institutional Ethics Committee, contact via Shamir Medical Center (Assaf Harofeh) Helsinki committee by for researchers who meet the criteria for access to confidential data.

## Data availability

Full sequencing data and reproducible analysis code will be released upon publishing.

